# Humoral and cellular responses to mRNA vaccines against SARS-CoV2 in patients with a history of CD20-B-cell depleting therapy

**DOI:** 10.1101/2021.07.04.21259848

**Authors:** Matthias B. Moor, Franziska Suter-Riniker, Michael P. Horn, Daniel Aeberli, Jennifer Amsler, Burkhard Möller, Linet M. Njue, Cesare Medri, Anne Angelillo-Scherrer, Luca Borradori, Susanne Radonjic-Hoesli, Morteza Seyed Jafari, Andrew Chan, Robert Hoepner, Vera Ulrike Bacher, Laila-Yasmin Mani, Joseena Mariam Iype, Cédric Hirzel, Britta Maurer, Daniel Sidler

**Affiliations:** Department of Nephrology and Hypertension, Inselspital, Bern University Hospital and Department of Biomedical Research, University of Bern, Bern, Switzerland; Department of Infectious Diseases, Inselspital, Bern University Hospital and University of Bern, Bern, Switzerland; Department of Clinical Chemistry, Inselspital University Hospital, Bern, Switzerland; Department of Rheumatology and Immunology, Inselspital, Bern University Hospital and Department of Biomedical Research, University of Bern, Bern, Switzerland; Department of Hematology and Central Hematology Laboratory, Inselspital, Bern University Hospital, University of Bern, Bern, Switzerland; Department of Dermatology, Inselspital, Bern University Hospital and University of Bern, Bern, Switzerland; Department of Neurology, Inselspital, Bern University Hospital and University of Bern, Bern, Switzerland

**Keywords:** Rituximab, CD20, B-cell depletion, Pfizer-BioNTech Comirnaty^®^, Moderna mRNA-1273, antibody, T-cells

## Abstract

**Background:** B-cell depleting therapies increase COVID19 morbidity and mortality. For this specific population, evidence-based vaccination strategies are lacking. Here, we investigated humoral and cell mediated immune responses to SARS-CoV2 mRNA-based vaccines in patients receiving CD20-B-cell depleting agents for autoimmune disease, malignancy, or transplantation.

**Methods:** Patients at the Bern University Hospital with a treatment history of anti-CD20 depleting agents (rituximab or ocrelizumab) were enrolled for analysis of humoral and cell-mediated immune responses (by interferon-γ release assay) after completing vaccination against SARS-CoV2. Primary outcome was the the anti-spike antibody response in anti-CD20-treated patients (n=96) in comparison to immunocompetent controls (n=29).

**Results:** Anti-spike IgG antibodies were detected in 49% of patients 1.79 months after the second vaccine dose (interquartile range, IQR: 1.16-2.48) compared to 100% of controls (p<0.001). SARS-CoV2 specific interferon-γ release was detected in 20% of patients and 75% of healthy controls (p<0.001). Only 11% of patients, but 75%of healthy controls showed positive reactions in both assays, respectively (p<0.001). Time since last anti-CD20 therapy (7.6 months), peripheral CD19+ (>27/µl), and CD4+ lymphocyte count (>653/µl) predicted humoral vaccine response (area under the curve [AUC]: 67% [CI 56-78], 67% [CI 55-80] and 66% [CI 54-79], (positive predictive value [PPV]: 0.78, 0.7 and 0.71).

**Conclusion:** This study provides evidence for blunted humoral and cell-mediated immune responses elicited by SARS-CoV2 mRNA vaccines in patients with CD20-depleting treatment history. Lymphocyte subpopulation counts are associated with vaccine response in this highly vulnerable population. (Funded by Bern University Hospital, ClinicalTrials.gov number, NCT04877496)

## Introduction

The coronavirus disease 2019 (COVID19) pandemic has taken a toll on many patients worldwide. Age and male sex are important drivers for severe COVID19 trajectories, but also preexisting autoimmune or kidney disease and malignancy ^1–3^. The backbone of pandemic-ending strategies is mass vaccination ^4,5^. Although randomized clinical trials of mRNA-based vaccines reported high vaccine efficacy^6^, these trials did not include immunocompromised patients, who can be expected to have inferior vaccination responses. This particularly applies to patients treated with B-cell depleting agents ^7^. Anti-CD20 B cell therapies are applied worldwide with annual doses ranging in the millions ^8^. In a broad spectrum of diseases with auto- and alloimmunity, and in hematological neoplasms, the B-cell depleting drug rituximab or biosimilar agents are serially administered. It has been shown that these patients are particularly vulnerable having 4x higher odds of COVID19-related mortality compared with those on other immunosuppressive medication such as methotrexate ^9^. The high variability of pharmacokinetics and thereby B cell recovery times ^10^, which makes the definition of ideal post treatment vaccination time points for mounting a sufficient immune response challenging. The need, however, for evidence-based recommendations, is large since in the absence of randomized, controlled trials, current recommendations to delay B-cell depleting therapies are based on previous influenza vaccination studies ^7^ and emerging humoral data on immune responses to SARS-CoV2 vaccines 11–16. An improved understanding of humoral and cell-mediated responses following SARS-COV2 mRNA vaccination in patients treated with anti-CD20 depleting agents is the prerequisite for the development of individualized vaccination strategies. Of note, recent data provided evidence that in COVID19, cell-mediated immune responses were crucial for vaccination efficacy 17–21 and might provide protection even in B cell depleted patients ^22^.

The lack of evidence-based vaccination strategies in this highly vulnerable population prompted the RituxiVac Study (NCT04877496), an investigator-initiated, single-center, case-control study to assess both humoral and cell-mediated immune responses to SARS-COV2 mRNA vaccines in patients with treatment history of anti-CD20 therapy.

## Methods

### Study design

The RituxiVac study was an investigator-initiated, single center, open-label, case-control trial conducted at the Departments of Nephrology and Hypertension, Rheumatology and Immunology, Hematology, Neurology, and Dermatology of the university hospital in Bern, Switzerland. COVID-naïve patients with a history of anti-CD20 therapy (rituximab or ocrelizumab) and completion of SARS-CoV2 vaccination for ≥4 weeks were enrolled between April 26 and June 30, 2021. Type, time points, cumulative dose and treatment indication for anti-CD20 therapies were recorded. All treatments since January 1, 2010 until the l date of first vaccination were considered. Additionally, age, gender and immunosuppressive medication were assessed. Type of vaccines and date of administration were derived from official records and COVID19 vaccination certificates. In addition, healthy controls without history of previous SARS-CoV-2 infection were enrolled at least four weeks after completion of their vaccine course.

Patients and healthy controls with prior SARS-CoV-2 infection were not eligible. All study participants were tested for the presence of anti-nucleocapsid antibodies. Individuals with positive results were excluded from the analysis. Individuals younger than 18 years of age and pregnant or lactating women were not eligible to participate.

In Switzerland, SARS-CoV2 vaccines were administered based on an age- and risk-tailored national priority plan. The BioNTech/Pfizer mRNA vaccine (Comirnaty®) was approved on December 19, 2020; the SARS-CoV2 vaccine Moderna® on January 12, 2021. All participants received two doses of either Comirnaty® or Moderna®. Allocation, administration and reporting of vaccination was coordinated and supervised by Swiss federal authorities independent of the study protocol.

The study was supported by internal institutional grants of the collaborators; the funders had no influence on the design or conduct of the trial and were not involved in data collection or analysis, in the writing of the manuscript, or in the decision to submit it for publication. The trial protocol was approved by the local ethics committee of the Canton of Bern, Switzerland (ID 2021-00669) and was registered on clinicaltrials.gov (Identifier: NCT04877496). The trial was performed in accordance with the principles of the Declaration of Helsinki, and all participants provided written informed consent prior to inclusion. The authors assume responsibility for the accuracy and completeness of the data and analyses, as well as for the fidelity of the trial and this report to the protocol.

### Study procedures

#### Baseline data collection

Trained study nurses and physicians completed a 17-item questionnaire for the post-vaccination study visit. Wherever available, dates and types of administered vaccines (Comirnaty® or Moderna®) were obtained from official vaccination records.

#### Blood collection and processing

For measurement of *IFN-γ release*, blood was collected in lithium heparin tubes (IFN-γ release assay) and serum tubes (antibody measurements, lymphocyte subpopulations).Serum tubes were centrifuged, and serum was then aliquoted and stored at -20°C prior to analyses. Creatinine values, lympocyte subpopulation counts, and total immunoglobulin quantities (IgG, IgM, IgA) were obtained using the routine clinical analytical services of the Center of Laboratory Medicine, Department of Clinical Chemistry of the University Hospital Bern.

#### anti-SARS-CoV2 S1-IgG and NC-IgG responses to vaccines

To assess humoral responses to vaccines, IgG antibodies targeting the SARS-CoV2 S1 protein, were detected using a commercial ELISA test from Euroimmun AG, Lübeck, Germany, as previously described 23. In brief, samples were diluted 1:100 in sample buffer. For antibody binding, 100 μL of diluted samples, prediluted positive and negative controls, and a prediluted calibrator were added for 1 hour at 37°C. After three washing steps, 100μl of HRP-labelled secondary anti-human IgG antibodies was added for 30 minutes at 37°C, followed by three more washing steps. Finally, 100 μL of TMB solution was added for 20 minutes. The reaction was stopped with 100 μL of 0.5M H2SO4, and results were measured at OD450-620 nm. Antibody values were expressed as ratio (ODsample/ODcalibrator). All samples with a ratio > 1.1 were considered as positive as per the manufacturer’s instructions. In order to exclude participants with previous COVID19, an anti-nucleocapsid ECLIA test was performed on a Cobas 8000 analyzer (Roche Diagnostics, Rotkreuz, Switzerland) ^24^. The cut-off was calculated based on the calibrator measurements and a cut-off index s/c ≥ 1.0 was considered positive as per the manufacturer’s instructions.

#### QuantiFERON® IFN-γ release responses to vaccine

To assess CMI responses to the vaccination, SARS-CoV2 specific interferon-γ release in whole blood was measured in a subset of participants (n=66) using QuantiFERON® SARS-CoV-2 Starter Pack (Qiagen Cat No./ID: 626715) that contains two different pools of protein S peptide. Following the manufacturer’s instructions, whole blood was incubated with peptide pools or mitogen for 1h. Subsequently, interferon-γ was quantified by ELISA (Qiagen Cat No./ID: 626410). All samples showed a positive response to mitogen. Responses to antigen pool 1 were analyzed. A cut-off value of 0.15 IU/ml was used to discriminate positive from negative CMI responses to SARS-CoV2, as reported before ^25^.

### Outcomes

The primary endpoint was the proportion of patients with a history of anti-CD20 treatment that showed a humoral immune response against SARS-CoV2 spike protein at least four weeks after completion of SARS-CoV2 vaccination, in comparison to immunocompetent controls. Humoral response was defined as anti-SARS-CoV2 S1 ≥ 1.1 (Index) ^26^.

Pre-specified secondary endpoints were the effect of anti-CD20 therapy, including time since last treatment and cumulative dose, on humoral or cell-mediated immune responses to SARS-CoV-2 mRNA-based vaccines in linear regression models adjusted for vaccine type, age, sex, immunosuppressive co-medication, and blood markers of immunocompetence (levels of IgG, IgM, IgA, absolute lymphocyte counts, absolute CD19- and CD3, CD4). After following the pre-specified data analysis, we assessed the discriminative power of selected biomarkers by calculating the area under the receiver operating characteristic curve (AUC-ROC) to predict vaccine elicited humoral and cell-mediated immune responses.

### Statistical analysis

Pre-screening revealed an eligible population of 725 participants with a history of at least one anti-CD20 treatment since January, 1^st^,.2010. With the assumption that 70% of patients and 98% of the healthy controls would reach the dichotomous outcome of humoral response, 18 healthy controls and 72 patients were determined as a minimal sample size with an enrollment ratio 4:1 (two-sided test, alpha error of 0.05, beta error of 0.8).

Statistical analyses were performed using R version 4.0.4 ^27^. A Chi-square test was used to compare categorical variables between two groups. Mann-Whitney U-test or t-test was used to compare continuous variables between groups, as appropriate. Linear regression analyses were obtained according to a plan using the lm function and logistic regression using the glm function in R. Selected regression models were visualized using the R package visreg ^28^. Area under the operator-received (AUC-ROC) curves were computed using package pROC. Statistical significance was determined at p<0.05. P values and widths of 95% confidence intervals have not been adjusted for multiplicity.

## Results

### Demographic and clinical characteristics of the patients

Overall, 106 patients and 30 healthy controls were enrolled in the RituxiVac study between April 26 and June 30, 2021 (Supplementary Figure 1). After exclusion of 6 participants (5 patients, 1 healthy control), who had positive anti-nucleocapsid and 5 patients, who received the first dose of rituximab after the first vaccine dose, our final study population consisted of 29 healthy controls and 96 patients. Anti-CD20 therapies were prescribed for autoimmune disease in 72 cases (75%), for malignancy in 7 cases (7.3%), and for induction therapy of ABO-incompatible kidney transplantation in 19 cases (20%). Demographic details, treatment history and vaccination data are presented in Table 1. Fifty-seven percent of patients reported immunosuppressive co-medication, among them corticosteroids in 44 cases (77%), calcineurin inhibitors in 19 cases (33%), antimetabolites in 24 cases (42%), methotrexate in 4 cases (7%), cytotoxic chemotherapy in 3 cases (5%), or other immunosuppressive drugs in 4 cases (7%). None of the healthy controls had treatment with h immunosuppressive agents or anti-CD20 therapy.

**Table 1:** Baseline characteristics, vaccination history of patients and healthy controls and anti-CD20-B-cell depletion history of patients in the study. Immunosuppression: any of prednisolone, calcineurin inhibitors, antimetabolites, methotrexate, cytotoxic chemotherapy or immunosuppressive/modulatory biologicals (apart anti-CD20). Median values (interquartile range) are given.

|  | **Patients**<br>**n = 96** | **Healthy Controls**<br>**n = 29** | **p-value** |
| --- | --- | --- | --- |
| **Male Sex - no of participants (%)** | 45 (47%) | 10 (34%) | 0.2 |
| **Median Age (IQR) - yr** | 67 (57, 72) | 54 (45, 62) | <0.001 |
| **Immunosuppression - no of participants (%)** | 57 (59%) | 0 (0%) | <0.001 |
| **Type of Vaccine (BioNTech/Pfizer) - no of participants (%)** | 58 (60%) | 9 (31%) | 0.005 |
| **Median Time since Vaccination (IQR) - months** | 1.79 (1.16, 2.48) | 1.81 (1.17, 2.48) | 0.5 |
| **Median Time since last anti-CD20 (IQR) - yr** | 1.07 (0.48, 2.55) | -- |  |
| **Median Cumulative Dose of anti-CD20 - (IQR) g** | 2.80 (1.50, 5.00) | -- |  |

### Anti-CD20 history and vaccination history

Patients were more frequently vaccinated with Comirnaty® manufactured by BioNTech/Pfizer(60%) versus controls (31%). Median time since last anti-CD20 treatment was 1.07 years (IQR: 0.48-2.55). 27%, 49% and 68% of patients received vaccination within 6, 12 and 24 months after last anti-CD20 therapy. Median cumulative dose of anti-CD20 depleting agent was 2.8 g (interquartile range, IQR: 1.5-5.0 g).

### Lymphocyte subpopulations and antibody levels

Baseline markers are provided in Table 2. Patients had significant lower peripheral CD3-, CD4- and CD19 cell counts (p<0.001). Absolute IgG and IgA levels were comparable, yet patients revealed moderately reduced IgM levels (p<0.01).

**Table 2:** Laboratory markers of immunocompetence in the peripheral blood of patients and healthy controls at time of study visit. Median values (interquartile range) are given. CD: cluster of differentiation, Ig: Immunoglobulin, µl: microliter, Reference: Normal Reference values.

|  | **Patients** | **Healthy Controls** |  |  |
| --- | --- | --- | --- | --- |
|  | **n = 96** | **n = 29** | **p-value** | **Reference** |
| **Lymphocytes (cells/µl)** | 1,344 (895, 1,720) | 2,275 (2,061, 2,368) | **<0.001** | 1200-2800 /µl |
| **CD3 Cells (cells/µl)** | 1,016 (662, 1,357) | 1,670 (1,312, 1,756) | **<0.001** | 690-2540 /µl |
| **CD4 Cells (cells/µl)** | 658 (459, 958) | 1,061 (958, 1,257) | **<0.001** | 410-1590 /µl |
| **CD19 Cells (cells/µl)** | 9 (1, 84) | 236 (228, 290) | **<0.001** | 90-660 /µl |
| **IgG (g/l)** | 8 (7, 10) | 10 (8, 10) | 0.14 | 7.0-16.0 g/l |
| **IgA (g/l)** | 1.62 (1.08, 2.41) | 1.74 (1.39, 2.34) | 0.5 | 0.7-4.0 g/l |
| **IgM (g/l)** | 0.48 (0.30, 0.73) | 0.80 (0.69, 1.22) | **<0.01** | 0.4-2.3 g/l |

### Vaccine elicited humoral and cell-mediated immune responses

We identified a median spike S1 IgG level of 7.34 (6.44-8.00) Index s/c in healthy controls and 0.74 (0.13-5.75) in anti-CD20 experienced patients (p<0.001) (Supplementary Table 1, Table 3). Anti-S1 IgG antibodies above the cut-off were present in 100% of healthy controls and 49% of patients (p<0.001). Stratified for treatment indication, the proportion of patients with positive anti-spike IgG antibodies treated for autoimmunity, ABO-incompatible transplantation and cancer was 60%, 40% and 11%, respectively. Similarly, cell-mediated immune responses to SARS-CoV-2 were significantly different among the groups, with SARS-CoV-2-specific IFN-γ release of 0.63 UI/ml (0.25-1.14) in healthy controls and 0.01 (0.00-0.08) in patients (p<0.0001). 75% of healthy controls and 17% of patients had therefore responses above the cut off provided by the manufacturer (p<0.001). Overall, 75% of healthy controls were double positive for anti-SARS-CoV2 spike IgG and cell-mediated response, whereas only 11% of patients showed this finding. Meanwhile, no healthy controls but 38% of patients remained double-negative. This uniformly demonstrates that a history of anti-CD20 therapy is associated with impaired humoral and cell-mediated responses induced by SARS-CoV2 mRNA vaccines.

**Table 3:** Frequency of positive humoral and cellular anti-SARS CoV2 response: Frequency of anti-SARS-CoV2 S1 IgG response above a threshold of 1.1 (Index s/c), interferon-gamma release above 0.15 IU/ml for patients and healthy controls.

|  | **Patients** | **Healthy Controls** |  |
| --- | --- | --- | --- |
|  | **n = 96** | **n = 29** | **p-value** |
| **Anti-SARS-CoV2 S1 IgG (>1.1 Index)** | 47/96 (49%) | 29/29 (100%) | **<0.001** |
| **IFN-γ release (>0.15 IU/ml)** | 13/66 (20%) | 21/28 (75%) | **<0.001** |
| **S1 IgG & IFN-γ positive** | 9/80 (11%) | 21/28 (75%) | **<0.001** |
| **S1 IgG & IFN-γ negative** | 31/82 (38%) | 0/29 (0%) | **<0.001** |

Finally, in an attempt to establish predictors for successful vaccination strategies in anti-CD20 treated patients, we analyzed demographics, medical history and biomarkers in linear regression models. In univariable models, age showed no effect on humoral responses, neither in patients nor in healthy controls (Figure 1A). However, peripheral CD19+ B-cell count (Figure 1B), total serum IgM levels (Figure 1C), time since last anti-CD20 treatment (Figure 1D), and CD4+ T-cell helper count (Figure 1E) showed positive associations with circulating anti-S antibodies (p<0.05). Total serum IgG levels were not associated with humoral vaccine responses (Figure 1F). In multivariable linear regression (Table 4), cumulative dose and time since last treatment were independent predictors for vaccine elicited humoral immune responses (p<0.001). Furthermore, Moderna® vaccine led to superior responses when compared to BioNTech/Pfizer Comirnaty® (p<0.01), while concomitant immunosuppressive medication independently blunted responses (p<0.001). Peripheral CD4+ T helper cell count, CD19+ cell count and IgM levels further independently predicted vaccination response. Peripheral CD4+ cell count and co-existing immunosuppression were the only determinants affecting vaccine elicited cell-mediated immune responses (p<0.05). Taken together, our analyses highlight the importance of anti-CD20 therapy timing and peripheral CD4+ and CD19+ lymphocyte counts, for immune responses to vaccines. To further explore these interactions, we plotted anti-SARS-CoV2 IgG levels against CD19 cell numbers and time delay since CD20-depletion for various levels of CD4 counts. Indeed, CD4 cells positively correlated in with IgG responses, notably in setting of low CD19 levels or short interval since CD20-B-cell depletion (Figure 1G-H).

**Table 4:** Multivariable linear regression for humoral and cellular anti-SARS CoV2 response. Multivariable linear regression of clinical and laboratory parameters important for immunocompetence in patients with history of anti-CD20 therapy. Interactions between the determinants was analyzed and given, if a p value <.05 was reached.

| | B coeff for anti-SARS-CoV2 IgG | | | B coeff for IFN-γ release | | |
| --- | --- | --- | --- | --- | --- | --- |
| | $\beta$ | 95% CI | p-value | $\beta$ | 95% CI | p-value |
| total Lymphocytes (per 1000/μl) | -1.10 | -3.2, 0.88 | 0.3 | 0.01 | -0.09, 0.11 | 0.9 |
| CD4 cells (per 1000/μl) | 4.50 | 1.3, 7.7 | **0.007** | -0.01 | -0.17, 0.16 | >0.9 |
| CD19 cells (per 1000/μl) | 10.00 | 4.6, 15 | **<0.001** | 0.64 | 0.26, 1.0 | **0.001** |
| IgG (per g/l) | -0.05 | -0.12, 0.02 | 0.13 | 0.00 | -0.01, 0.01 | 0.4 |
| IgG (per g/l) | 0.24 | -0.34, 0.81 | 0.4 | -0.03 | -0.08, 0.02 | 0.2 |
| IgG (per g/l) | 1.40 | 0.49, 2.2 | **0.003** | -0.02 | -0.07, 0.04 | 0.5 |
| | $\beta$ | 95% CI | p-value | $\beta$ | 95% CI | p-value |
| Male Sex (yes) | -0.23 | -1.4, 0.94 | 0.7 | -0.14 | -0.32, 0.03 | 0.1 |
| Age (per year) | -0.01 | -0.06, 0.03 | 0.6 | -0.01 | -0.01, 0.00 | 0.13 |
| Cum. anti-CD20 Dose (per g) | 0.33 | 0.14, 0.52 | **<0.001** | -0.01 | -0.04, 0.02 | 0.5 |
| Time since anti-CD20 (per year) | 0.55 | 0.24, 0.86 | **<0.001** | -0.01 | -0.05, 0.04 | 0.8 |
| Immunosuppression (yes) | -2.10 | -3.3, -0.93 | **<0.001** | -0.21 | -0.38, -0.04 | **0.017** |
| Vaccine Type (Moderna<sup>®</sup>) | 1.70 | 0.53, 2.9 | **0.005** | 0.10 | -0.07, 0.27 | 0.3 |

**Figure 1:**
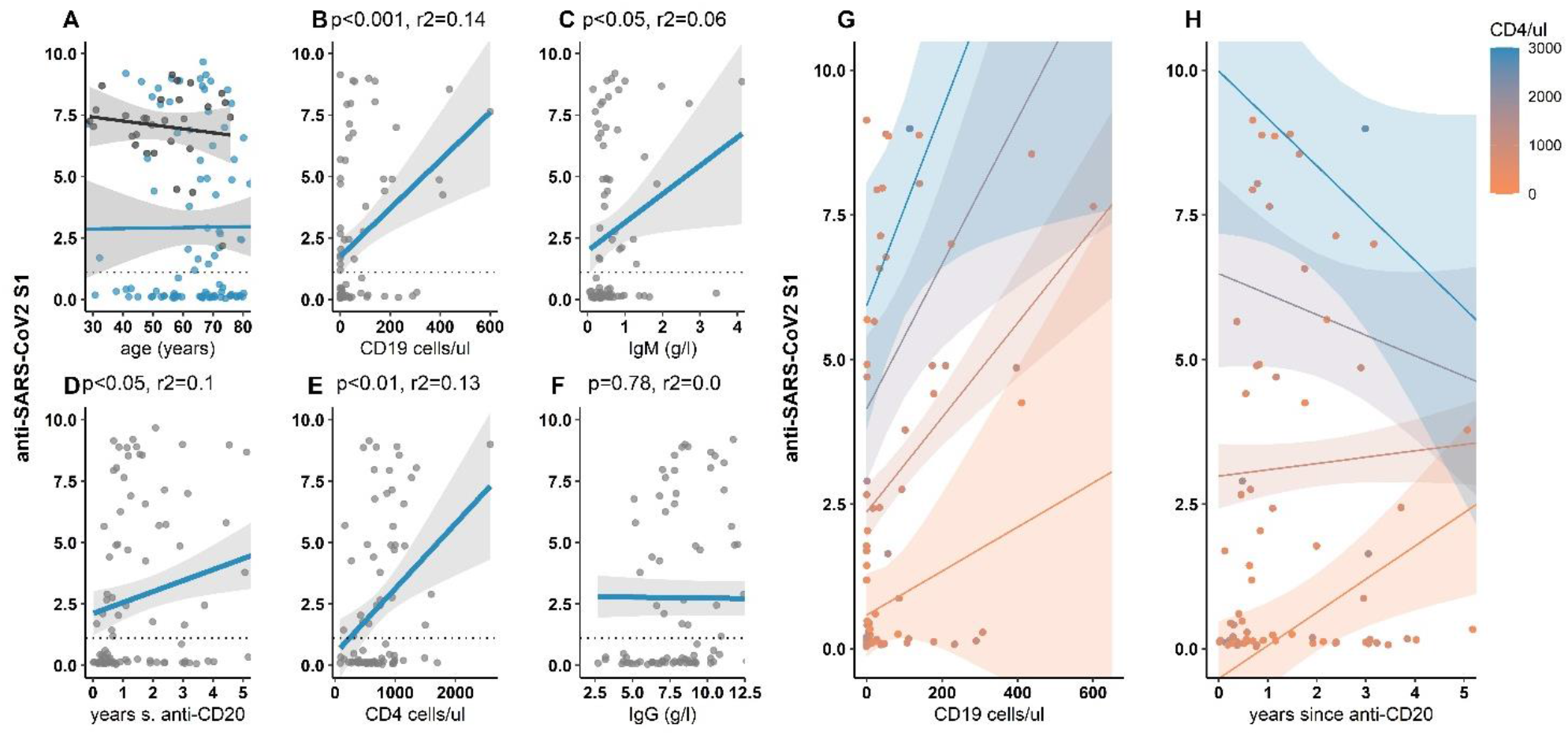
Univariate Correlation between anti-SARS-CoV2 S1-IgG and clinical and serological parameters of immunocompetence. (A). Linear regression between anti-SARS-CoV2 S1-IgG levels and participant’s age. Black: Healthy controls, blue: patients. (B-F): Linear regression between anti-SARS-CoV2 S1-IgG levels and indicated parameters, blue: Linear regression for given parameters for patients. Shaded ribbon: 95% CI for regression line. grey: individual values. Each point represents one patient. r2 represents the regression coefficient. Dotted line: cutoff anti-SARS-CoV2 S1-IgG value of 1.1 (s/c). (G, H): Linear regression between anti-SARS-CoV2 S1-IgG levels and CD19 cell count (G) and time since CD20-depletion (years) (H) for various levels of CD4 cell count.

Next, we applied ROC curves to evaluate the classification performance of the three most promising clinical and laboratory characteristics to predict humoral response to SARS-CoV2 vaccines. IgG levels, CD19+ and CD4+ cell counts are surrogate markers for immune competence. Figure 2 shows sensitivity and specificity of time since last treatment, peripheral CD19+ count and CD4+ count to predict a dichotomous anti-SARS-CoV2 humoral response. All three showed significant divergence from the null diagonal with similar area under the curve (66-67%). Analyses revealed optimal cutoffs at 0.64 years since last treatment, 27 CD19+ cells/µL and 653 CD4+ cells/µL with respective positive predictive values of 0.78, 0.70 and 0.71.

**Figure 2:**
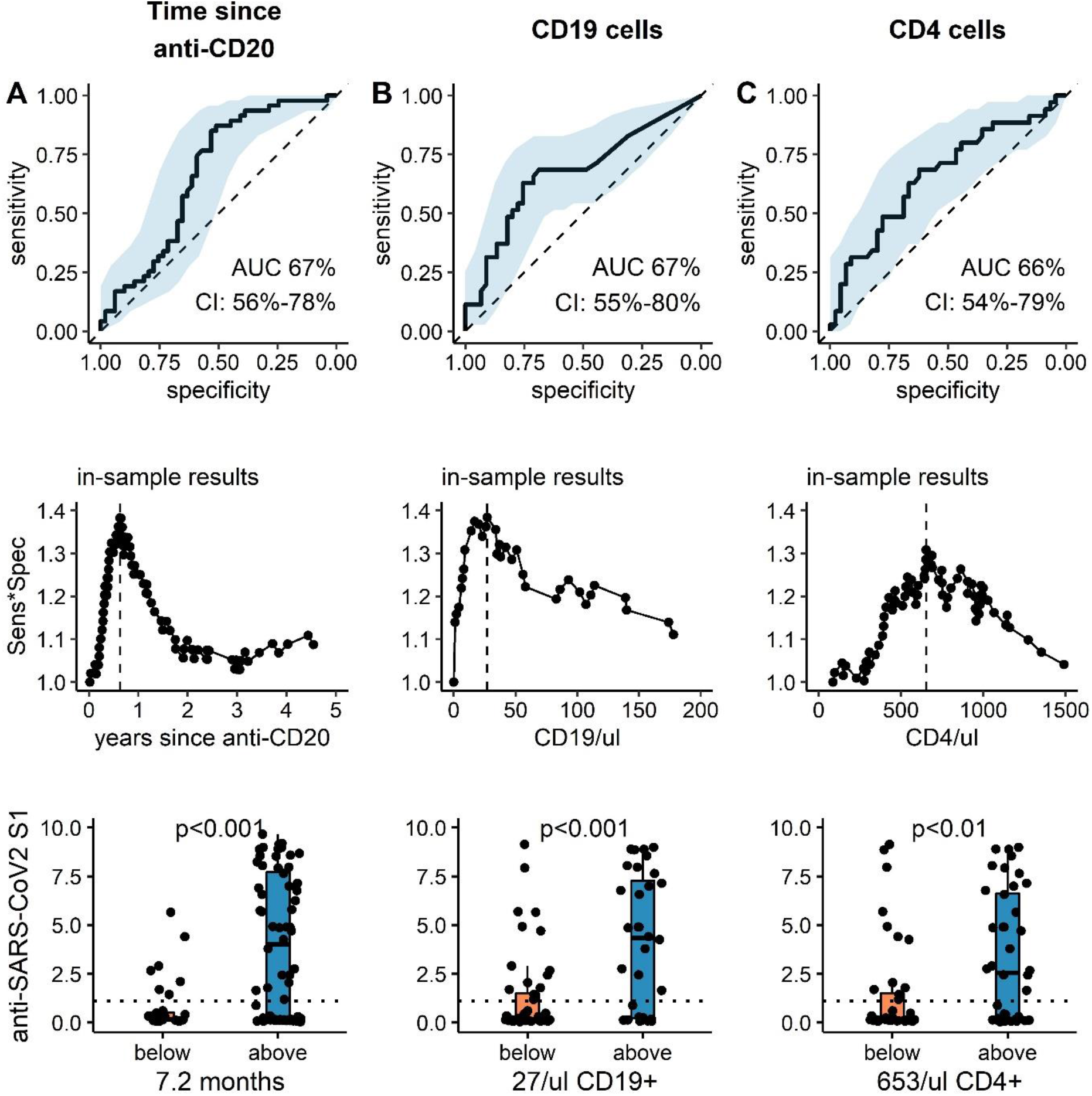
Receiver operating characteristic curve, optimized cut-off and predictive value of clinical and serological parameters to predict anti-SARS-CoV2 humoral response. (top panel): ROC curve for (A) time since last anti-CD20 treatment, (B) CD19 count and (C) CD4 count to predict dichotomous anti-SARS-CoV2 S1 IgG levels above 1.1 (Index s/c) at least 4 weeks after the second COVID vaccine. Black solid line: ROC curve, blue ribbon: 95% CI, dotted line: null hypothesis. (middle panel): Calculation of optimal cutoffs to predict dichotomous anti-SARS-CoV2 S1 IgG response using Youden method. Product of Sensitivity and Specificity is plotted against continuous variable: (A) time since CD20-depletion (years), (B) CD19 count and (C) CD4 count. Lower panel: Absolute anti-SARS-CoV2 S1 IgG are shown (median, IQR and min/max). For patients with parameters below (orange) or above (blue) the respective cutoff values. Additionally, individual values of each patient are plotted as single points. Dotted line: cutoff anti-SARS-CoV2 -IgG value of 1.1 (s/c). p-values: *<0.05, **<0.01.

In summary, ROC curves of treatment history or generic markers for immune competence, namely CD4+ and CD19+ cell count reveal cutoff points to classify humoral responders and non-responders, which can guide future studies directly addressing biomarkers of vaccination responses in larger populations.

To foster these results, we analyzed peripheral immune cell counts and vaccine responses depending on time since last anti-CD20 treatment. As expected, CD19+ B cell counts rose with longer delay since last treatment: At <6 months 9%, at <12 months 20%, and >12 months after anti-CD20 treatment 62% of patients fulfilled the threshold of 27 CD19+ B cells/µl, suggestive of favorable vaccination outcome. Meanwhile, for the same time points, 50%, 44% and 56% of patients met the criterion of at least 653 CD4 cells/µl, which has similar predictive value for successful SARS-CoV2 vaccination (Supplementary Table 2). Humoral responses were low in the first 6 months after anti-CD20 therapy and increased thereafter. Meanwhile, cell-mediated was stable for all patient groups, independent on time since anti-CD20 treatment. In the subgroup of patients with less than 6 months between anti-CD20 therapy and vaccination, a CD4 cutoff of 653 µl/l had a positive predictive value of 0.9.

## Discussion

Our investigator-initiated, single center, open-label, case-control trial in 125 individuals shows that interval since last CD20-depletion and levels of circulatory CD19 and/or CD4 cells predict SARS-CoV2 vaccination response in patients with a history of anti-CD20 therapy. These parameters could thus be used to optimize and individualize vaccination strategies.

Two factors made the present study uniquely suited to evaluating the immune responses to SARS-COV2-mRNA-vaccines in a real-life setting. First, a rare combination of detailed clinical and laboratory background data provided by the integrative data repository center of our tertiary referral center at the university hospital Bern and second, the rapid pace of the vaccination campaign with a correspondingly short post-vaccination follow-up period.

Rituximab and biosimilars are critical backbones in the treatment of patients with autoimmunity and/or B-cell mediated malignancy. A sufficiently frequent anti-CD20 dosing and suppression of peripheral B-cells are treatment goals in such patients, notably those with active and/or progressive disease. Therefore, a tailored vaccination strategy based on CD20-depletion interval or peripheral B-cell count would deem many patients ineligible for vaccination, namely those with the most aggressive treatment regimens, which are at greatest risk for severe COVID19 trajectories.

Our report adds novelty to the results of a series of smaller studies that have previously investigated humoral and cellular responses to SARS-CoV2 vaccination of patients with a history of B-cell depleting anti-CD20 therapies 11,12,16,29,30.

In our study, humoral responses against SARS-CoV2-specific mRNA vaccines were observed in all healthy controls, yet only in 49% of anti-CD20 treated patients. When stratified for the treatment indication, patients with autoimmune diseases had a higher response rate as compared with post-transplantation or cancer patients, which might arise from differences in concomitant immunosuppressive treatment. Similarly, vaccine elicited cellular responses occurred in 86% of healthy controls and 17% of patients. Overall, whereas the majority of healthy individuals mounted both, successful humoral and cellular vaccination responses, this only applied to 5% of anti-CD20 exposed patients thereby underlining the complex sequelae of B cell depletion on B and T cell interactions. In addition to the expected lower CD19+ B cell counts and IgM levels, patients showed decreased numbers of CD3+ and CD4+ T cells. These findings support the notion that selective B-cell depletion indirectly results in a reduction of certain subsets of T-lymphocytes 31,32, which may further impair vaccination efficacy.

Strengths of this study include the identification of potential predictors for vaccination efficacy in anti-CD20 treated patients. For adequate humoral responses, timing of anti-CD20 therapy, CD19+ counts, and IgM levels were crucial. Interestingly, CD4+ counts positively predicted both adequate humoral and cellular responses. These results as well as the observed positive correlation of CD4+ cell counts with anti-Spike IgG antibodies support an important role for T cells for vaccination efficacy in B cell depleted patients as recently suggested 30. Interestingly, although the COVID-19 Vaccine Moderna® and Corminaty® (BioNTech/Pfizer) are very similar apart from differences in the structure of the lipid nanoparticles 33, Moderna® prompted both superior humoral and cellular responses. Another important observation was the fact that immunosuppressive co-medication impaired both humoral and cellular vaccine elicited immune responses, which is an important point to consider for the individualization of vaccination strategies. Cut-off points for CD4+ and CD19+ counts defined by ROC analysis allowed to distinguish vaccination responders from non-responders at a given time interval after last anti-CD20 therapy.

The present study has limitations. First, we could not measure anti-SARS-CoV2 spike protein antibodies before vaccination, because the Swiss vaccination program accelerated in high-risk individuals before recruitment was initiated. Therefore, we measured currently circulating anti-SARS-CoV2 nucleocapsid antibodies and excluded the three participants with detectable anti-nucleocapsid antibodies. Second, the distribution of vaccines manufactured by Pfizer-BioNTech and Moderna was not identical between controls and patients. Comirnaty® was the earliest approved vaccine in Switzerland that was made primarily available for individuals at high risk for severe COVID19. By the time when health care professionals and the general population became eligible for SARS-COV2 vaccination, Moderna® constituted the largest share of vaccines delivered to Switzerland. Finally, the current study population was highly heterogeneous regarding the underlying diseases, indications for CD20 depletion treatment, and immunosuppressive co-medication. However, this complex study population represents a real-world scenario and provided a unique opportunity to explore routine laboratory parameters that are readily available and could be used to predict vaccination efficacy and therefore guide vaccination timing despite the complexity of the various immunosuppressive regimens.

Based on the current data, we propose that a simple peripheral count of CD4+ cells could serve as a starting point to stratify patients according to anticipated vaccination response, even in patients with recent and/or severe B-cell depletion. Given the nowadays broad availability of SARS-CoV2 vaccines and the good tolerability, vaccination should be offered to all patients, if success can be predicted with acceptable certainty.

To conclude, the present data establish a severely impaired humoral and cellular response to SARS-CoV2 mRNA vaccines in patients with a history of B-cell depleting anti-CD20 therapies including rituximab and ocrelizumab. Our analyses provide first estimates of ideal peripheral CD19+ and CD4+ cell counts and time since last dose of anti-CD20 therapy that allow a positive humoral response to SARS-CoV2 vaccines. Upon validation in independent cohorts in a prospective setting, these results may provide a critical guide for coordinating both the administration of vaccines and B-cell depleting agents in this population.

## Data Availability

Data are available from the authors on request.

## Acknowledgements

The authors wish to thank all volunteers and patients for their participation, all involved nurses and study nurses including Barbara Strehler, Sabine Hasler, Mark Wienand and Ruth Kober for their contribution to data collection. The authors thank statistician Andreas Limacher, PhD of the Clinical Trials Unit, University of Bern for advice on the statistical analysis plan. Furthermore, the authors thank Monika Hurni, Juliette Schlatter, Olivier Schaerer, Thomas Mormot and Rodoljub Pavlovic for technical assistance.

## Data availability

Data are available from the authors on request.

## Authors’ contributions

MBM,BM and DS conceived the study. MBM, FRS, MPH, CH, BM and DS designed the study. MBM, DA, BM, LN, CM, AAS, LB, SRH, MSJ, AC, RH, VUB, LYM, BM and DS recruited participants. MBM and DS performed computational chart review. MBM, FRS, MPH, CH and BM have verified the underlying data and performed analyses. MBM, BM and DS wrote the manuscript with input from all authors. All authors approved the final version of the manuscript.

## Funding

The current study was funded by Bern University Hospital. The funder had no role in design, interpretation of results, manuscript drafting or decision to publish the results.

**Supplementary Figure 1:**
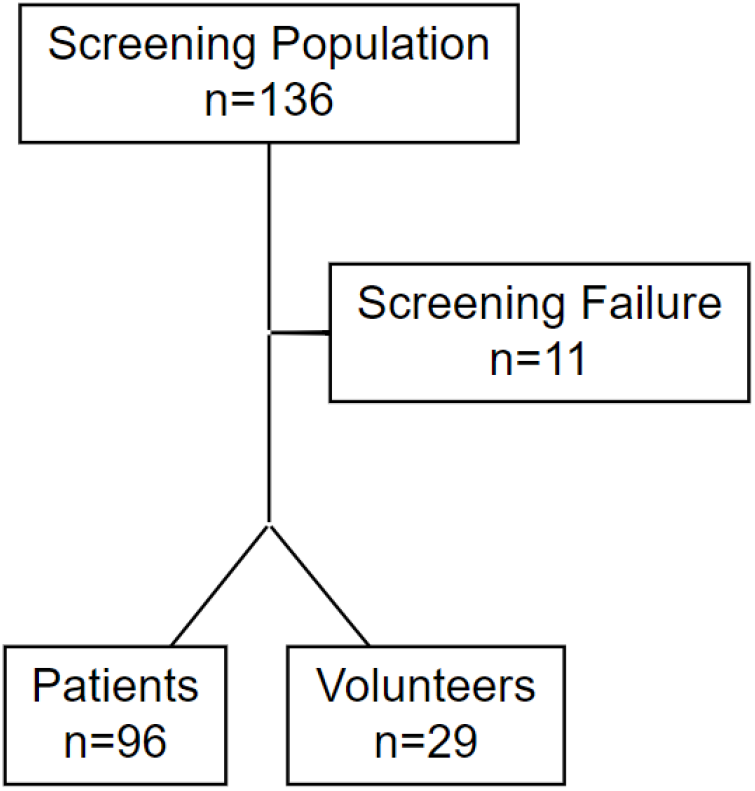
Flowchart of screening and grouping of patients and healthy volunteers in the RituxiVac study.

**Supplementary Table 1:** Absolute humoral and T-cellular anti-SARS CoV2 response: anti-SARS-CoV2 S1 levels in the plasma and interferon γ-concentration from peripheral blood mononuclear cells stimulated with a SARS-CoV2 spike peptide pool. Median values (interquartile range) are given.

|  | **Patients** | **Healthy Controls** | **p-value** |
| --- | --- | --- | --- |
| **anti SARS-CoV2 S1 (Index)** | 0.74 (0.13, 5.75) | 7.34 (6.44, 8.00) | **<0.001** |
| **IFNγ Release (IU/ml)** | 0.01 (0.00, 0.08) | 0.52 (0.16, 0.86) | **<0.001** |

**Supplementary Table 2:**
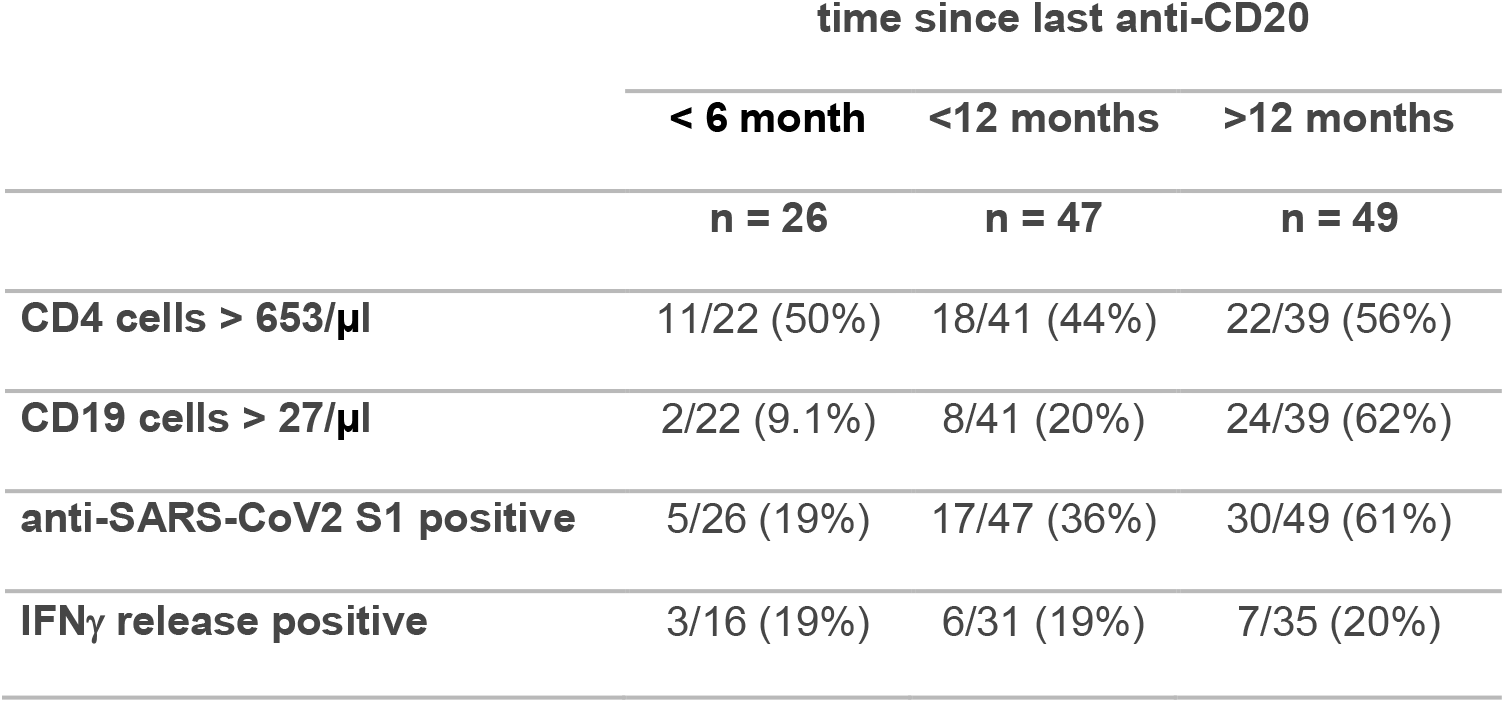
CD4, CD19 cell count and humoral and T-cellular anti-SARS CoV2 response in patients in anti-CD20 treated patients: 26, 47 and 49 patients had a treament intervall between last anti-CD20 dose and vaccination of <6, <12 and >12 months, respectively. Fraction of patients, who fulfilled the CD4 cutoff (>653/µl) or CD19 cutoff (>27/µl) and the study visit and fraction of patients who with anti-SARS-CoV2 S1 above a threshold of 1.1 (Index s/c) and interferon-gamma release above 0.15 IU/ml.

